# Effectiveness of the Influenza Vaccine During the 2024-2025 Respiratory Viral Season: A Prospective Cohort Study

**DOI:** 10.1101/2025.01.30.25321421

**Authors:** Nabin K. Shrestha, Patrick C. Burke, Amy S. Nowacki, Steven M. Gordon

## Abstract

**Background:** The purpose of this study was to evaluate the effectiveness of the influenza vaccine during the 2024-2025 respiratory viral season.

**Methods:** Employees of Cleveland Clinic in employment in Ohio on October 1, 2024, were included. The cumulative incidence of influenza among those in the vaccinated and unvaccinated states was compared over the following 33 weeks. Protection provided by vaccination (analyzed as a time-dependent covariate) was evaluated using multivariable Cox proportional hazards regression, including adjusting for influenza activity as an effect modifier.

**Results:** Among 53402 employees, 43920 (82.2%) were vaccinated by the end of the study. Influenza occurred in 1130 (2.12%) during the study. The cumulative incidence of influenza was similar for the vaccinated and unvaccinated states early, but over time the cumulative incidence of influenza increased more rapidly among the vaccinated than the unvaccinated. In a multivariable Cox proportional hazards regression analysis, the risk of influenza was not significantly different during periods when influenza activity was low (HR, .70; 95% C.I., .42 - 1.15; *P* = .15) or medium (HR, 1.20; 95% C.I., .86 - 1.67; *P* = .28), but was significantly higher for the vaccinated state than the unvaccinated state when influenza activity was high (HR, 1.33; 95% C.I., 1.07 - 1.64; *P* = .009).

**Conclusions:** This study was unable to find a protective influence of influenza vaccination among working-aged adults during the 2024-2025 respiratory viral season and found that influenza vaccination was associated with a higher risk of influenza when influenza activity was high.

**Summary:** Among 53402 working-aged Cleveland Clinic employees, we were unable to find a protective influence of influenza vaccination during the 2024-2025 respiratory viral season and found a significantly higher risk of influenza with vaccination when influenza activity was high.

## INTRODUCTION

Influenza is a common respiratory viral infection with potential for substantial mortality and morbidity, and was estimated to be responsible for 145 000 deaths worldwide among all ages in 2017 [1]. While seasonal outbreaks are typical, predominantly occurring over the winter months [2], influenza also has the potential to cause pandemics with far greater impact, as exemplified by the 1918 pandemic, which was estimated to have a case fatality rate of 2.5% and resulted in an estimated 50 million deaths worldwide [3]. The virus evolves continuously through antigenic drift, leading to seasonal strain variation, and can occasionally undergo more dramatic changes through antigenic shift, which may give rise to pandemics [4]. This ongoing viral evolution underscores the importance of effective and timely vaccination strategies.

Influenza is also a vaccine-preventable illness. However, antibody responses following influenza vaccination rise within weeks but decline substantially over subsequent months, often returning to near baseline by 6–12 months post-vaccination [5,6], and annual influenza vaccination is recommended at the beginning of each respiratory viral season in the autumn months in the northern hemisphere [7].

Additionally, the effectiveness of the vaccine in any given year depends on how similar the strains contained in the vaccine are to the strains causing infection that year. The most widely used seasonal influenza vaccine is the trivalent inactivated vaccine (TIV), which is composed of two influenza A virus types (H3N2 and H1N1) and an influenza B virus type [7,8]. A new vaccine is produced each year in an attempt to match the vaccine strains to the strains projected to be most prominent in the upcoming influenza season. Since the current process of developing the vaccine typically takes a few months, a decision on which strains to include in the vaccine must be made several months in advance. In years where there is a good match between the vaccine strains and the infecting strain, vaccine effectiveness is expected to be good. In years where there is a poor match between vaccine strains and the circulating infecting strain, vaccine effectiveness is expected to be poor.

Given the high morbidity and mortality burden of influenza, universal annual vaccination against the infection is recommended by the Advisory Committee on Immunization Practices [7]. Over the last couple of decades, policies of mandatory annual vaccination of healthcare personnel have been adopted increasingly across healthcare institutions [9].

Healthcare resource utilization, including hospitalizations, and resource needs such as quantity of antiviral medications needed, are strongly affected by how effective the vaccine is during any respiratory viral season. Early estimates of vaccine effectiveness of the influenza vaccine during any respiratory viral season can provide information that can help healthcare institutions and pharmacies prepare for the remainder of the season.

The Centers for Disease Control and Prevention (CDC) has a network to estimate vaccine effectiveness (VE) in the United States each year and has published interim vaccine effectiveness estimates for the 2024-25 season. The study reported the VE to be surprisingly good when considering that practicing physicians in our part of the country observed that a substantial proportion of vaccinated persons acquired influenza during this season, and it was not uncommon for entire vaccinated families to have the infection, thereby raising serious questions about the accuracy of the VE estimates obtained.

The purpose of this study was to evaluate the effectiveness of the influenza vaccine during the 2024-2025 respiratory viral season in North America.

## METHODS

### Study design

This was a prospective cohort study conducted at the Cleveland Clinic Health System (CCHS) in the United States.

### Patient Consent Statement

The study was approved by the Cleveland Clinic Institutional Review Board as exempt research (IRB no. 23-625). A waiver of informed consent and waiver of HIPAA authorization were approved to allow the research team access to the required data.

### Setting

For several years Cleveland Clinic has had a mandatory participation influenza vaccination program, which requires employees to either receive an annual influenza vaccine or seek an exemption on medical or religious grounds. The vaccine is provided to healthcare personnel free of charge. Vaccines received outside CCHS are actively recorded in the occupational health database. When healthcare personnel develop acute respiratory illnesses, they are encouraged to seek medical attention and the decision to test for influenza is made on a case-by-case basis by the treating provider either in the occupational health clinics or at their personal providers’ offices.

### Participants

CCHS employees in employment at any Cleveland Clinic location in Ohio on the study start date were included in the study. Individuals with missing data on age or sex were excluded from the analysis (3% of the cohort). Both age and sex are known to be associated with both the likelihood of influenza vaccination and the risk of influenza infection, and were therefore considered essential covariates for adjustment. Given the small proportion of missing data and the importance of these variables, we opted for a complete case analysis rather than imputation, as the potential for bias from exclusion was considered minimal.

### Variables

Data were obtained from the institution’s electronic data vault, which serves as a centralized repository of electronic health record data. Variables collected were influenza vaccination date, age, sex, job location, job type (categorized as clinical nursing or other), and date of positive test for influenza. Job type was dichotomized in this way to isolate the group with consistently high levels of direct patient contact, primarily bedside nurses, whose intensity of direct patient contact may put them at elevated occupational risk for influenza exposure. The “other” category encompassed a broad range of roles, including but not limited to physicians, respiratory therapists, medical assistants, lab personnel, administrative staff, and environmental services. Institutional data governance around employee data limited our ability to collect additional clinical variables.

Influenza was defined as a positive nucleic acid amplification test for influenza A or B any time after the study start date. Only molecular (including molecular point-of-care tests) performed within Cleveland Clinic Health System were included.

### Outcome

The study outcome was time to influenza. Outcomes were followed until May 21, 2025.

### Statistical analysis

For the 2024-2025 influenza season, the vaccine became available on 1 October 2024. This date was considered the study start date.

To assess potential differences in the propensity for influenza testing between vaccinated and unvaccinated individuals, we calculated, for each day of the study period, the ratio of the proportion of vaccinated individuals who underwent PCR testing for influenza to the proportion of unvaccinated individuals similarly tested. Additionally, we examined the ratio of the proportion of influenza PCR tests positive among the vaccinated versus the unvaccinated on each day of the study. These analyses allowed us to evaluate whether differential testing behavior or differences in test positivity by vaccination status might bias vaccine effectiveness estimates.

A Simon-Makuch hazard plot [10] was created to compare the cumulative incidence of influenza in the vaccinated and unvaccinated states, by treating influenza vaccination as a time-dependent covariate [11,12]. Individuals who had not developed influenza were censored at the end of the study follow-up period. Those whose employment was terminated during the study period before they had influenza were censored on the date of termination of employment. Only first episodes of influenza were included in the analysis. Curves for the unvaccinated state were based on data while the vaccination status of individuals remained “unvaccinated”. Curves for the vaccinated state were based on data from the date the influenza vaccination status changed to “vaccinated”.

We conducted time-to-event analyses using Cox proportional hazards regression to estimate the association between vaccination status and the risk of influenza infection. Vaccination status was modeled as a time-dependent covariate, changing from unvaccinated to vaccinated 7 days following the date of vaccination to allow for time to develop vaccine-induced immunity. Influenza activity in the population was categorized into three levels based on quantiles of influenza activity (“low” for quantiles 1-3, “medium” for quantile 4, and “high” for quantile 5), and included as an effect modifier. The primary model included an interaction term between vaccination status and influenza activity level, and adjusted for age, sex, clinical nursing job, and primary work location. This allowed estimation of hazard ratios (HRs) for vaccination status within each influenza activity stratum.

Hazard ratios and 95% confidence intervals (CIs) for vaccination status at low influenza activity were obtained directly from the model coefficients. For medium and high influenza activity levels, combined effect estimates were calculated by summing the coefficient of vaccination status and the corresponding interaction term. Variances and covariances of these coefficients were used to compute accurate confidence intervals and p-values for these combined estimates.

Variance inflation factors were evaluated to ensure that there was no multicollinearity in the models. The proportional hazards assumption was checked by examining Schoenfeld residuals and there was no indication of violations.

Statistical significance was assessed at the 0.05 level. The analysis was performed by N. K. S. and A. S. N. using the *survival* package and R version 4.4.2 (R Foundation for Statistical Computing) [13].

## RESULTS

A total of 53402 employees in Ohio remained after excluding 1700 individuals (3.1%) for whom age or gender were missing. These employees formed the study cohort and a total of 43920 (82.2%) were vaccinated by the end of the study. The vaccine was the inactivated trivalent influenza vaccine in 98.6% of those vaccinated. Altogether, 1130 employees (2.12%) acquired influenza during the 33 weeks of the study. Of these, 1109 (98.1%) were influenza A infections, the remaining being influenza B infections. A total of 3679 individuals (6.9%) were censored during the study period because of termination of employment before the end of the study. The duration of person-time follow up was 2,934,076 days in the unvaccinated state, and 8,920,343 days in the vaccinated state.

Influenza activity in our region began to rise in mid-November, peaked in late December to early January, and declined by early March, which aligns with typical seasonal patterns in our area. Ninety-eight percent of influenza cases detected among study participants were influenza A, with subtyping (where available) indicating that the predominant circulating strain was influenza A (H1N1). This distribution closely mirrored community-wide influenza surveillance data in our region during the same period.

### Baseline characteristics

Table 1 shows the characteristics of individuals included in the study. Notably, this was a relatively young population, with a mean age of 42 years, and 75% were female. Twenty percent had a clinical nursing job.

**Table 1.** Baseline characteristics of 53402 employees of Cleveland Clinic in Ohio.

| Characteristics | Overall<sup>a</sup> |
| --- | --- |
| Age in years, mean (SD) | 42.0 (13.4) |
| Sex |  |
| Female | 40 130 (75.1) |
| Male | 13 272 (24.9) |
| Primary work location |  |
| Cleveland Clinic Main | 20 536 (38.5) |
| Regional hospitals<sup>b</sup> | 21 880 (41.0) |
| Ambulatory centers | 9351 (17.5) |
| Administrative centers | 1635 (3.1) |
| Job type |  |
| Clinical nursing job | 10 840 (20.3) |
| Not clinical nursing job | 42562 (79.7) |

<sup>a</sup>Data are presented as no. (%) unless otherwise indicated.
<sup>b</sup>Includes Akron General, Ashtabula, Euclid, Fairview, Hillcrest, Lodi Community, Lutheran, Marymount, Medina, Mentor, Mercy (Canton), Southpointe, and Union, Hospitals, all part of the Cleveland Clinic Health System.

### Testing differences between the vaccinated and unvaccinated

The ratio of the proportion of the vaccinated who underwent PCR testing for influenza to the proportion of the unvaccinated similarly tested on each day of the study was significantly higher than 1.00 for most of the study period (Figure 1), particularly during the period when a large number of cases of influenza were being diagnosed, suggesting that the vaccinated were more likely to be tested than the unvaccinated on any given day.

**Figure 1.**
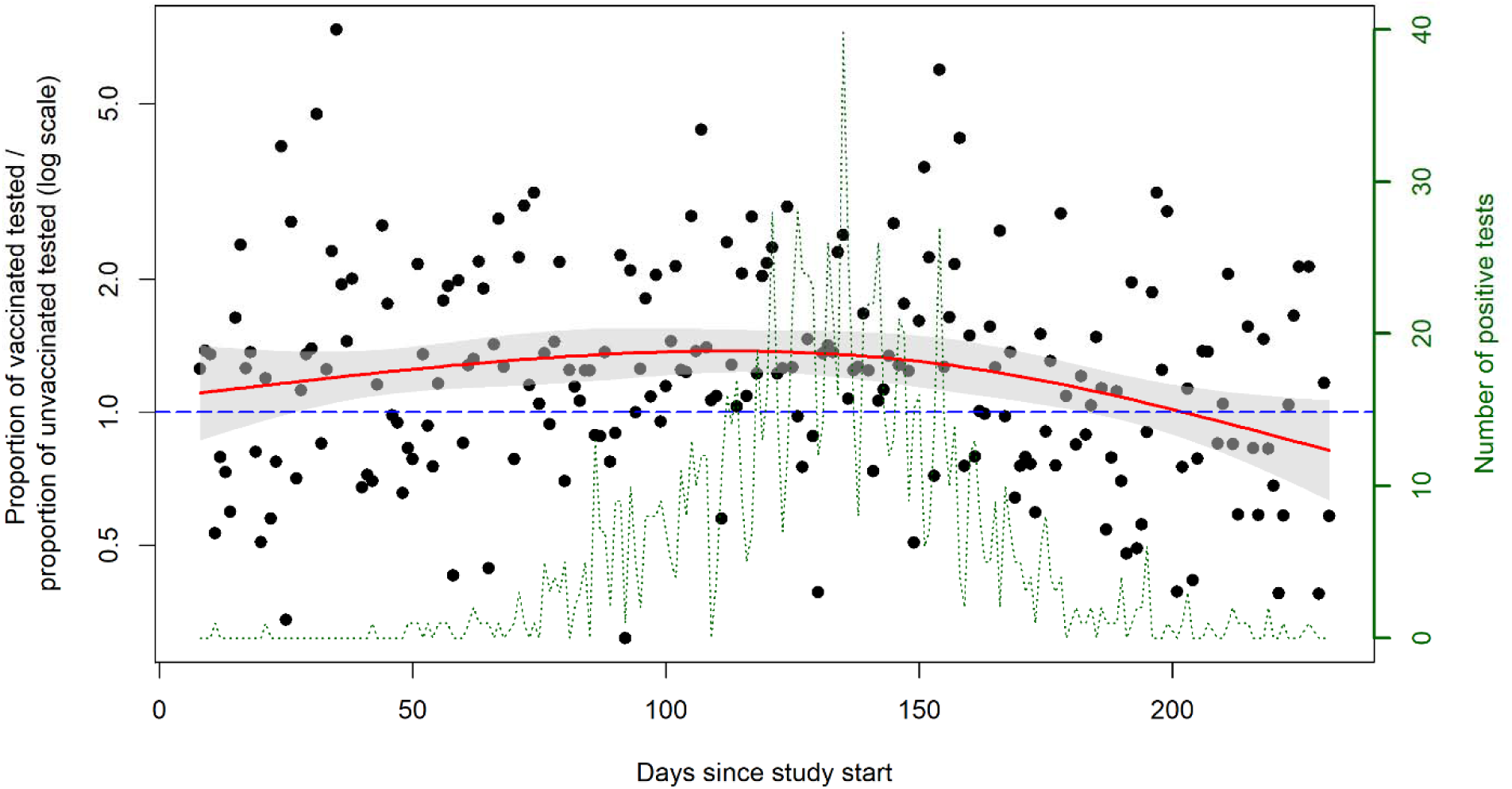
Comparison of the ratio of the proportion of the vaccinated who underwent PCR testing for influenza to the proportion of the unvaccinated who underwent PCR testing for influenza on each day of the study. The y-axis is represented on a logarithmic scale. Each day is represented by a dot. The dashed line represents the reference line where the testing proportions are the same for those vaccinated and unvaccinated. Dots representing days on which a higher proportion of vaccinated than unvaccinated individuals were tested for influenza will fall above the reference line, and dots for days on which a lower proportion of vaccinated than unvaccinated individuals were tested for influenza will fall below the reference line. The red line represents the best fit line for the geometric mean of the above ratio by restricted cubic spline regression, after excluding outliers (values >3 standard deviations from the mean ratio), with the shaded areas representing its 95% confidence interval. The dotted green line is a trend plot showing the daily number of positive influenza tests over the study period. **Alt text:** Figure comparing the proportion of the vaccinated who underwent PCR testing for influenza to the proportion of the unvaccinated who underwent PCR testing for influenza, on each day of the study

During the period when most infections occurred (days 110 – 170, when 74% of all the positive influenza tests occurred), the ratio of proportion of tests positive among the vaccinated to that among the unvaccinated on each day of the study was not significantly different from 1.00 (Figure 2). This finding suggested that the increased testing observed among the vaccinated may primarily reflect a higher number of infections rather than a greater propensity to seek testing. This analysis indicated that if there was a difference in healthcare-seeking behavior between the vaccinated and the unvaccinated, such was not detected in our data.

**Figure 2.**
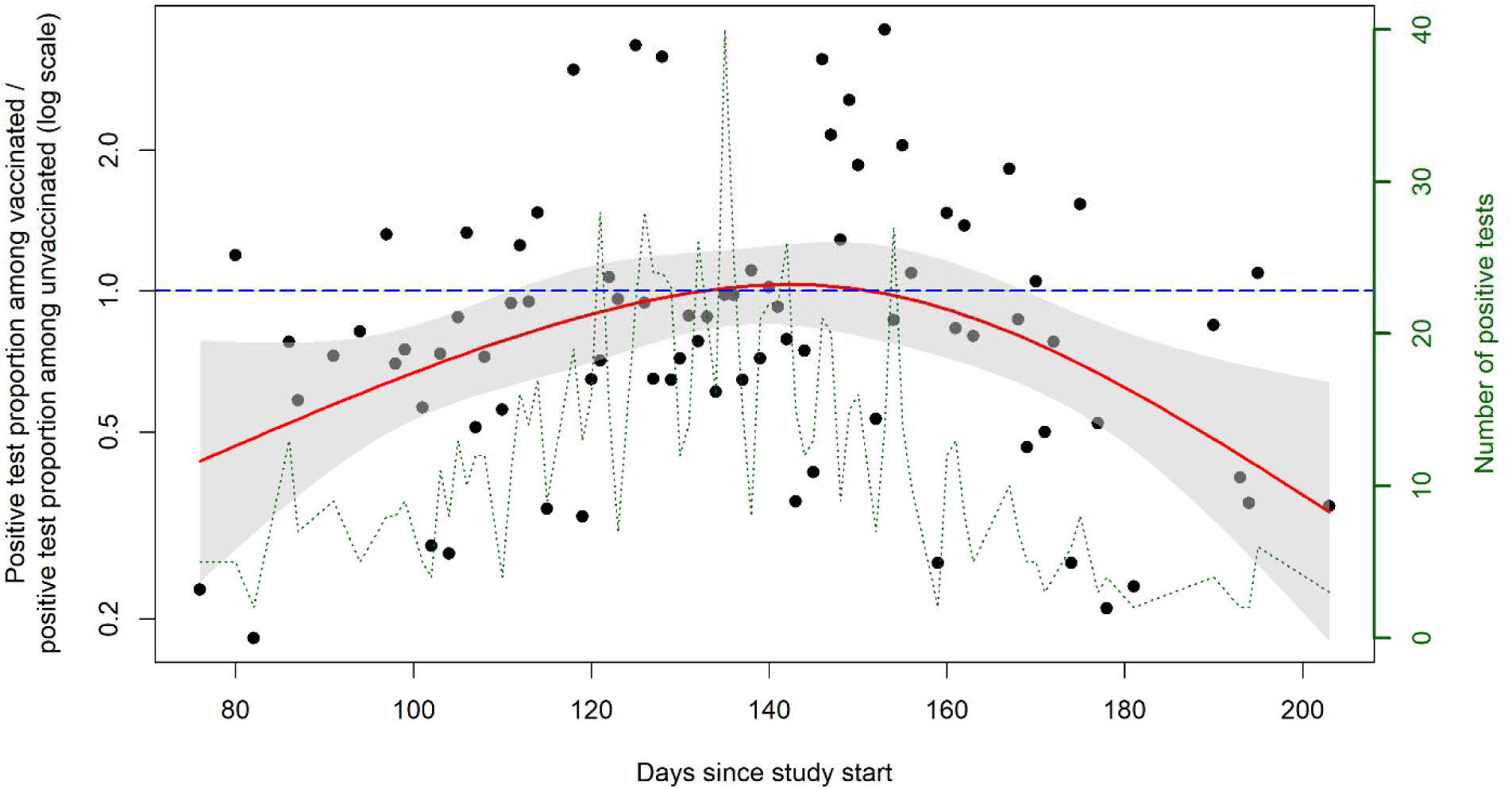
Comparison of the ratio of the proportion of vaccinated persons’ tests that were positive to the proportion of unvaccinated persons’ tests that were positive on each day of the study. Each day is represented by a black dot. The blue dashed line represents the reference line where the proportion of tests positive are the same for those vaccinated and unvaccinated. Dots representing days on which the vaccinated had a higher proportion of tests positive than the unvaccinated will fall above the reference line, and dots for days on which the vaccinated had a lower proportion of tests positive than the unvaccinated will fall below the reference line. The red solid line represents the best fit line for the geometric mean of the above ratio by restricted cubic spline regression, after excluding outliers (values >3 standard deviations from the mean ratio), with the shaded areas representing its 95% confidence interval. This was based on data for days where both vaccinated and unvaccinated had at least one test done. Data were inadequate to obtain data points prior to day 76 of the study. The dotted green line is a trend plot showing the daily number of positive influenza tests over the study period. **Alt text:** Figure comparing the proportion of vaccinated persons’ tests that were positive to the proportion of unvaccinated persons’ tests that were positive, on each day of the study.

### Influenza vaccine effectiveness

Very few individuals developed influenza A in the first two months of the study and the daily number of infections began to increase steadily about 70 days after the study start date. The cumulative incidence of influenza did not appear to be significantly different between the vaccinated and unvaccinated states early on, but over the course of the study the cumulative incidence of infection increased more rapidly among the vaccinated than the unvaccinated (Figure 3). In a multivariable Cox proportional hazards regression analysis adjusted for age, sex, clinical nursing job, and primary employment location, and allowing for influenza activity to be an effect modifier, the risk of influenza did not differ significantly for the vaccinated compared to the unvaccinated state during periods of low influenza activity (HR, .70; 95% C.I., .42 - 1.15; *P* = .15) or medium influenza activity (HR, 1.20; 95% C.I., .86 - 1.67; *P* = .28), but was significantly higher for the vaccinated state than the unvaccinated state when influenza activity was high (HR, 1.33; 95% C.I., 1.07 - 1.64; *P* = .009). Point estimates and 95% confidence intervals for hazard ratios for acquisition of influenza, for the various variables in unadjusted and adjusted Cox proportional hazards regression models, are shown in Table 2.

**Figure 3.**
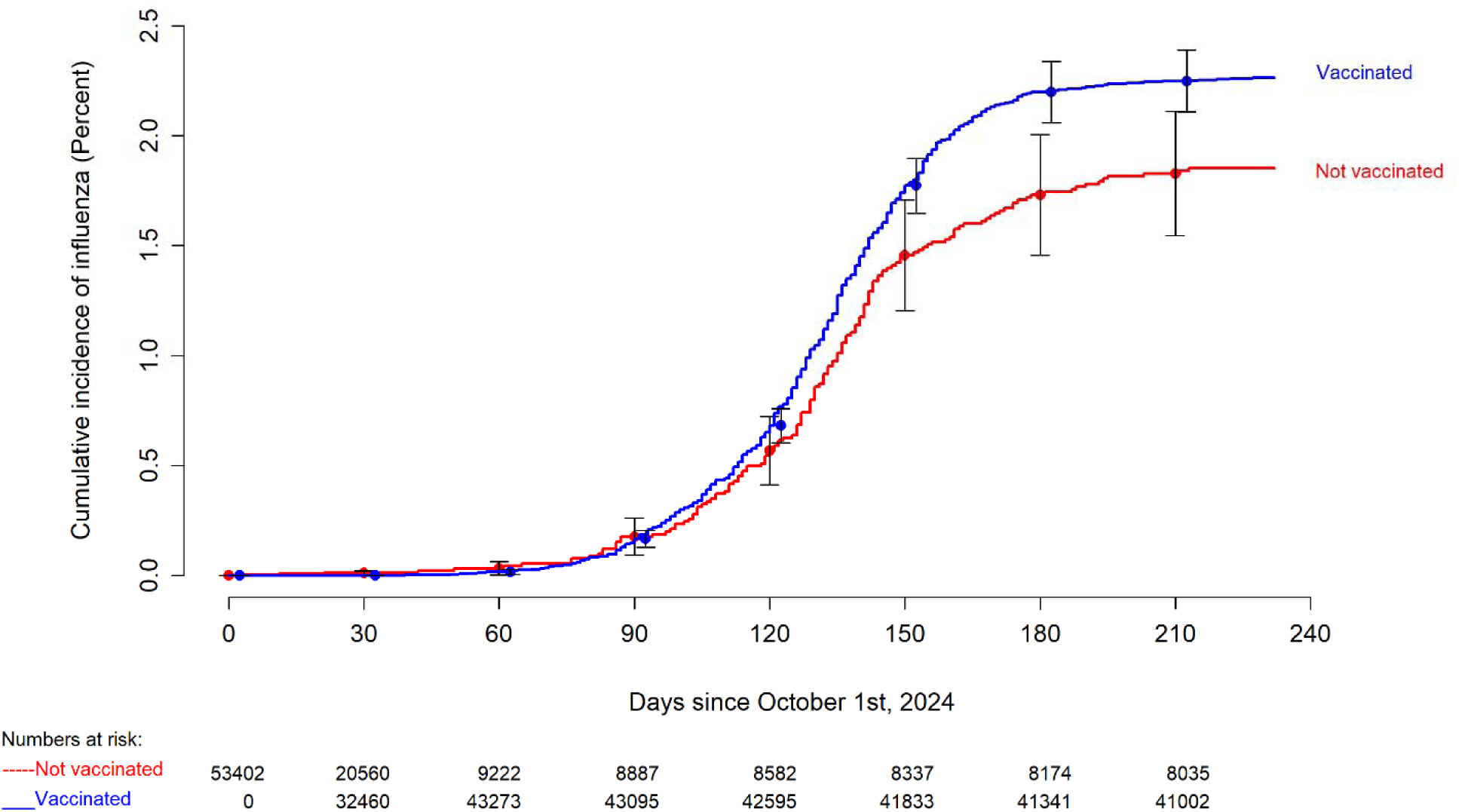
Simon-Makuch plot comparing the cumulative incidence of influenza for individuals stratified by vaccination status. Day zero was 1 October 2024, the day the influenza vaccine began to be offered to employees for the respiratory viral season. Point estimates and 95% confidence intervals are jittered along the x-axis to improve visibility. **Alt text:** Figure comparing the cumulative incidence of influenza among the vaccinated and the unvaccinated.

**Table 2.**
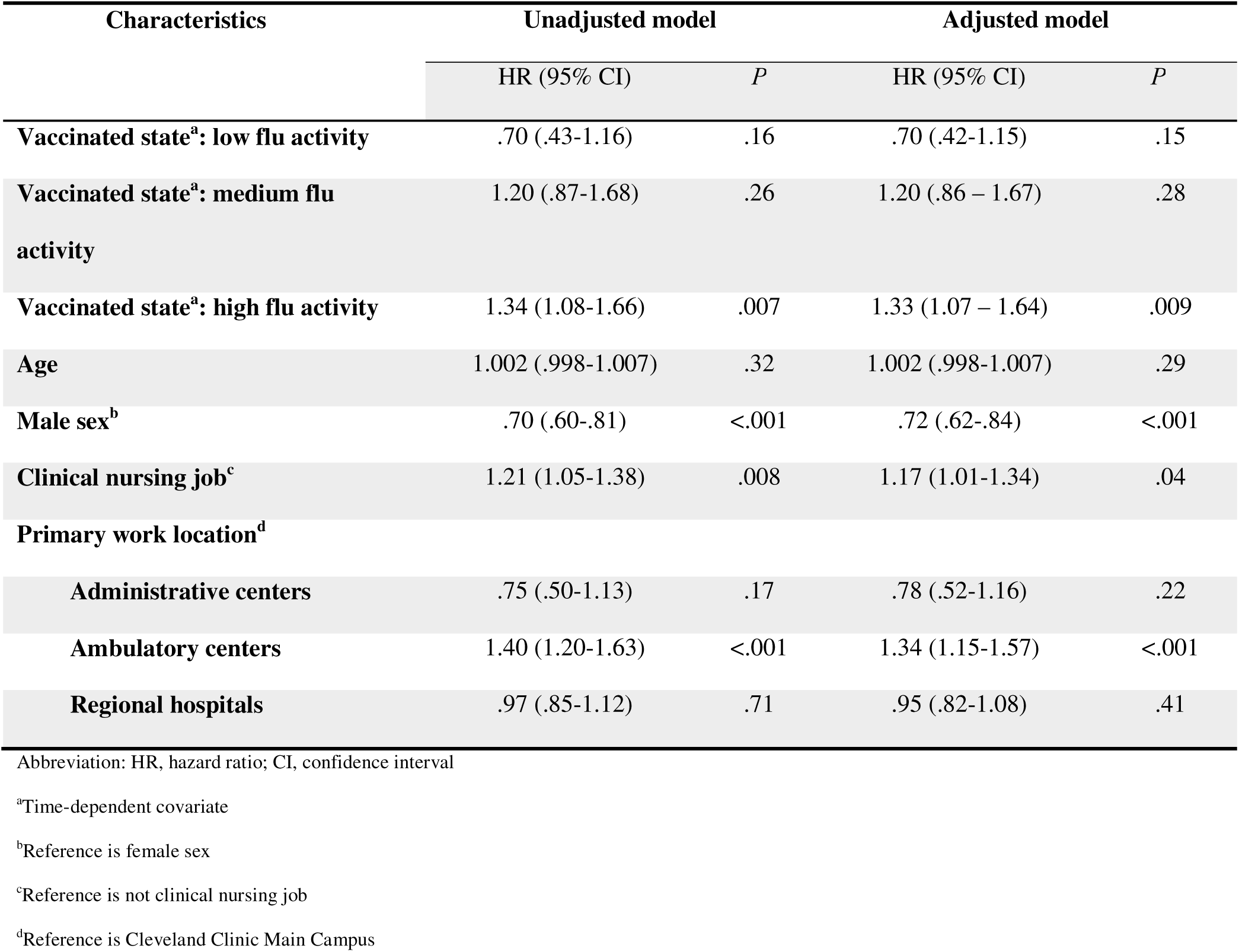
Unadjusted and Adjusted Associations with Time to Influenza in Cox Proportional Hazards Regression Models.

## DISCUSSION

This study found a significantly higher risk of influenza among the vaccinated compared to the unvaccinated state in northern Ohio during the high influenza activity period of the 2024-2025 influenza season. These findings contrast with the CDC’s interim estimates of influenza vaccine effectiveness for the same season, which suggested moderate protection against medically attended influenza in both outpatient and inpatient settings, based on a test-negative design study [14].

There are possible explanations for this discrepancy. Test-negative studies estimate VE by comparing the odds of vaccination among individuals who test positive for influenza to those who test negative, under the assumption that vaccinated and unvaccinated individuals have similar propensities to seek care and testing. However, in real-world settings this assumption may not hold. Vaccinated individuals may be more likely to seek testing for milder symptoms due to heightened health awareness. This differential healthcare-seeking behavior could lead to an overrepresentation of vaccinated individuals among those testing negative, thereby inflating VE estimates.

Our study mitigated this concern by using a cohort design with vaccination modeled as a time-dependent covariate, allowing for direct estimation of relative risk rather than extrapolating from odds ratios. While test-negative design studies can approximate rate ratios when controls are selected concurrently with cases (i.e., incidence density sampling) [15], these odds ratios are often interpreted as risk ratios in VE calculations, which may misrepresent vaccine performance when the vaccinated and unvaccinated have different propensities to seek testing, particularly during high-incidence periods. Our study design provides a more intuitive and transparent understanding of the vaccine’s performance in a real-world population. Although our study design was still limited by the possibility of differential propensities to get tested among the vaccinated and the unvaccinated, it allowed for such differences to be analyzed. Limitations of the available data did not permit direct adjustment for a person’s propensity to get tested. However, we found that although vaccinated individuals were more likely to undergo PCR testing for influenza than unvaccinated individuals (suggesting the potential for confounding), the proportion of positive tests was not significantly different between groups when most cases of influenza occurred (providing some reassurance that the difference in testing was not from difference in propensity to get tested). This suggests that the increased testing among the vaccinated was likely due to a higher number of infections rather than increased testing behavior alone. Since such an analysis cannot completely rule out residual confounding due to healthcare-seeking behavior, we performed a sensitivity analysis to assess how robust our primary finding was to potential unmeasured or uncontrolled confounding, by calculating the evidence value (E-value) [16,17]. The E-value assesses the impact of a missing confounder on an observed effect. We found the E-value to be 1.99, suggested that an unmeasured confounder would need to have a HR of 1.99, conditional on the measured covariates, with both vaccination status and the outcome of influenza, to explain away the significant association of increased risk of influenza with vaccination during high influenza activity (HR = 1.33), making it unlikely that a missing confounder could completely explain the observed association. Additionally, our use of a time-dependent covariate approach to vaccination status allowed for determining vaccine effectiveness in real time, which provided us with very early signals about the magnitude of vaccine effectiveness within a few weeks of the first cases of influenza being diagnosed.

Our study also has limitations. Nearly all participants received the trivalent inactivated influenza vaccine; therefore, the results may not generalize to other influenza vaccines, including LAIV. Infections diagnosed via home testing would not have been captured. A potential limitation is that individuals vaccinated outside the system may also seek influenza testing elsewhere when ill. While their vaccination status is recorded through the mandatory reporting process, test results from outside facilities may not be captured. This would have the effect of disproportionately undercounting influenza cases among those classified as vaccinated, because they would be classified as vaccinated but their outcome would be no influenza, potentially leading to an overestimation of vaccine effectiveness. The study was not powered to assess rare outcomes such as influenza-associated hospitalizations or mortality, nor was it designed to evaluate vaccine impact on illness severity. Furthermore, our cohort consisted of working-age healthcare personnel, limiting generalizability to children, the elderly, and severely immunocompromised populations. Despite these limitations, our findings provide important real-world data on influenza vaccine effectiveness in a key target population. The results are generalizable to relatively healthy adults in the USA, which is a major target of adult influenza vaccination efforts. Although the study was done in northern Ohio, there is little reason to assume that the effectiveness of the vaccine would have been different in a different geographic region within the continental USA.

Although the increased risk of influenza with influenza vaccination appears to be counter-intuitive, there is biological plausibility for why this could possibly happen. Antigenic imprinting refers to a phenomenon where the first exposure of the immune system to influenza by infection or vaccination shapes the breadth of immune responses to subsequent influenza infections or vaccinations, preferentially recalling memory B cells targeting epitopes of the originally encountered strain rather than generating new responses to current strains [18–20]. A study examining the impact of repeated influenza vaccinations on vaccine effectiveness over 8 seasons in Wisconsin found significantly higher vaccine effectiveness among individuals with no prior vaccine history compared to individuals frequently vaccinated [21]. A study in Canada evaluating the impact of prior vaccination in one or two preceding seasons found, that compared to those unvaccinated all three seasons, those vaccinated in the current season only or in the current season and only one prior season had a lower risk of influenza, those vaccinated in one of the prior seasons only had a similar risk of influenza, and those vaccinated in all three seasons had a significantly higher risk of influenza [22]. There have also been studies that have not found a negative effect of prior vaccination [23,24]. It has been suggested that the effect of prior vaccination may depend on the antigenic relatedness of the previous vaccine to the current vaccine, and of both to the currently circulating virus, a concept referred to as the antigenic distance hypothesis [25]. These studies suggest a need for a deeper understanding of the effects of prior vaccination so that inhibitory effects of prior vaccination can be mitigated.

Given all the variables that can influence the effectiveness of the influenza vaccine in any given year, and our current processes for developing the vaccine, one should not expect the vaccine to be highly effective year after year. It therefore becomes important to evaluate the effectiveness of the vaccine every year. This study’s inability to find a protective influence of influenza among adults in the healthcare workforce in northern Ohio, USA, during the 2024-2025 winter season, and indeed an increased risk of influenza associated with influenza vaccination during high influenza activity, demonstrates a need to develop systems to examine influenza vaccine effectiveness early during each respiratory season and be open to making adjustments to vaccine policy accordingly.

## Data Availability

All data produced in the present study are available upon reasonable request to the authors

## Notes

### Author contributions

N. K. S.: Conceptualization, methodology, validation, investigation, data curation, software, formal analysis, visualization, writing-original draft preparation, writing-reviewing and editing, supervision, project administration. P. C. B.: Resources, investigation, validation, writing-reviewing and editing. A. S. N.: Methodology, formal analysis, visualization, validation, writing-reviewing and editing. S. M. G.: Resources, writing-reviewing and editing.

